# Integrated host-microbe metagenomics for sepsis diagnosis in critically ill adults

**DOI:** 10.1101/2022.07.16.22277700

**Authors:** Katrina Kalantar, Lucile Neyton, Mazin Abdelghany, Eran Mick, Alejandra Jauregui, Saharai Caldera, Paula Hayakawa Serpa, Rajani Ghale, Jack Albright, Aartik Sarma, Alexandra Tsitsiklis, Aleksandra Leligdowicz, Stephanie Christenson, Kathleen Liu, Kirsten Kangelaris, Carolyn Hendrickson, Pratik Sinha, Antonio Gomez, Norma Neff, Angela Pisco, Sarah Doernberg, Joseph L. Derisi, Michael A. Matthay, Carolyn S. Calfee, Charles R. Langelier

## Abstract

Sepsis is a leading cause of death, and improved approaches for disease diagnosis and detection of etiologic pathogens are urgently needed. Here, we carried out integrated host and pathogen metagenomic next generation sequencing (mNGS) of whole blood (n=221) and plasma RNA and DNA (n=138) from critically ill patients following hospital admission. We assigned patients into sepsis groups based on clinical and microbiological criteria: 1) sepsis with bloodstream infection (Sepsis^BSI^), 2) sepsis with peripheral site infection but not bloodstream infection (Sepsis^non-BSI^), 3) suspected sepsis with negative clinical microbiological testing; 4) no evidence of infection (No-Sepsis), and 5) indeterminant sepsis status. From whole blood gene expression data, we first trained a bagged support vector machine (bSVM) classifier to distinguish Sepsis^BSI^ and Sepsis^non-BSI^ patients from No-Sepsis patients, using 75% of the cohort. This classifier performed with an area under the receiver operating characteristic curve (AUC) of 0.81 in the training set (75% of cohort) and an AUC of 0.82 in a held-out validation set (25% of cohort). Surprisingly, we found that plasma RNA also yielded a biologically relevant transcriptional signature of sepsis which included several genes previously reported as sepsis biomarkers (e.g., *HLA-DRA, CD-177*). A bSVM classifier for sepsis diagnosis trained on RNA gene expression data performed with an AUC of 0.97 in the training set and an AUC of 0.77 in a held-out validation set. We subsequently assessed the pathogen-detection performance of DNA and RNA mNGS by comparing against a practical reference standard of clinical bacterial culture and respiratory viral PCR. We found that sensitivity varied based on site of infection and pathogen, with an overall sensitivity of 83%, and a per-pathogen sensitivity of 100% for several key sepsis pathogens including *S. aureus, E. coli, K. pneumoniae* and *P. aeruginosa*. Pathogenic bacteria were also identified in 10/37 (27%) of patients in the No-Sepsis group. To improve detection of sepsis due to viral infections, we developed a secondary RNA host transcriptomic classifier which performed with an AUC of 0.94 in the training set and an AUC of 0.96 in the validation set. Finally, we combined host and microbial features to develop a proof-of-concept integrated sepsis diagnostic model that identified 72/73 (99%) of microbiologically confirmed sepsis cases, and predicted sepsis in 14/19 (74%) of suspected, and 8/9 (89%) of indeterminate sepsis cases. In summary, our findings suggest that integrating host transcriptional profiling and broad-range metagenomic pathogen detection from nucleic acid may hold promise as a tool for sepsis diagnosis.

## Introduction

Sepsis causes 20% of all deaths globally and contributes to 20-50% of hospital deaths in the United States alone^1, 2^. Early diagnosis and identification of the underlying microbial pathogens is essential for timely and appropriate antibiotic therapy, which is critical for sepsis survival^3, 4^. Yet in over 30% of cases, no etiologic pathogen is identified^5^, reflecting the limitations of current culture-based microbiologic diagnostics^6^. Adding additional complexity is the need to differentiate sepsis effectively from non-infectious systemic illnesses, which often appear clinically similar at the time of hospital admission.

As a result, antibiotic treatment often remains empiric rather than pathogen-targeted, with clinical decision-making based on epidemiological information rather than individual patient data. Similarly, clinicians often continue empiric antimicrobials despite negative microbiologic testing for fear of harming patients in the setting of falsely negative results. Both scenarios lead to antimicrobial overuse and misuse, which contributes to treatment failures, opportunistic infections such as *C. difficile* colitis, and the emergence of drug-resistant organisms^7^.

With the introduction of culture-independent methods such as metagenomic next generation sequencing (mNGS), limitations in sepsis diagnostics may be overcome^8, 9^. Recent advancements in plasma cell-free DNA sequencing have expanded the scope of metagenomic diagnostics by enabling minimally invasive detection of circulating pathogen nucleic acid originating from diverse anatomical sites of infection^9^. The clinical impact of plasma DNA metagenomics has been questioned, however, due to frequent identification of microbes of uncertain clinical significance, inability to detect RNA viruses that cause pneumonia, and limited utility in ruling-out presence of infection^10, 11^.

Whole blood transcriptional profiling offers the potential to mitigate these limitations by capturing host gene expression signatures that distinguish infectious from non-infectious conditions, and viral from bacterial infections^12, 13^. However, because transcriptional profiling exclusively captures the host response to infection, it does not provide precise taxonomic identification of sepsis pathogens, which limits the utility of this approach when performed alone. Further, transcriptional profiling has traditionally required isolating peripheral-blood mononuclear cells, or stabilizing whole blood in specialized collection tubes, and it has remained unknown whether a simple plasma specimen could yield informative data for host-based infectious disease diagnosis.

In recent work, a single-sample metagenomic approach combining host transcriptional profiling with unbiased pathogen detection was developed to improve lower respiratory tract infection diagnosis^14^. Sepsis, defined as, “life-threatening organ dysfunction from a dysregulated host response to infection^15^,” provides an additional clear use case for this integrated host-microbe metagenomics approach. Here, we study a prospective cohort of critically ill adults to develop a novel sepsis diagnostic assay that combines host transcriptional profiling with broad-range pathogen identification. By applying machine learning to high dimensional mNGS data, we evaluate host and microbial features that distinguish microbiologically confirmed sepsis from non-infectious critical illness. We then demonstrate that plasma nucleic acid can be used to profile both host and microbe for precision sepsis diagnosis.

## Results

### Clinical features of study cohort

We conducted a prospective observational study of critically ill adults admitted from the Emergency Department (ED) to the Intensive Care Unit (ICU) at two tertiary care hospitals (**Figure 1**). Patients were categorized into five subgroups based on sepsis status (**Methods**). These included patients with: 1) clinically adjudicated sepsis and a microbiologically confirmed bacterial bloodstream infection (Sepsis^BSI^), 2) clinically adjudicated sepsis and a microbiologically confirmed non-bloodstream infection (Sepsis^non-BSI^), 3) suspected sepsis with negative clinical microbiologic testing (Sepsis^suspected^), 4) patients with no evidence of sepsis and a clear alternative explanation for their critical illness (No-Sepsis), or 5) patients of indeterminant status (Indeterm). The most common diagnoses in the No-Sepsis group were cardiac arrest, overdose/poisoning, heart failure exacerbation, and pulmonary embolism. The majority of patients, regardless of subgroup, required mechanical ventilation and vasopressor support (**Supplementary Table 1**). Patients with microbiologically proven sepsis (Sepsis^BSI^ + Sepsis^non-BSI^) did not differ from No-Sepsis patients in terms of age, gender, race, ethnicity, immunocompromise, APACHEIII score, maximum white blood cell count, intubation status, or 28-day mortality (**Figure 1**, **Supplementary Table 1**). All but one patient (in the No-Sepsis group) exhibited ≥ 2 systemic inflammatory response syndrome (SIRS) criteria^16^.

**Figure 1.**
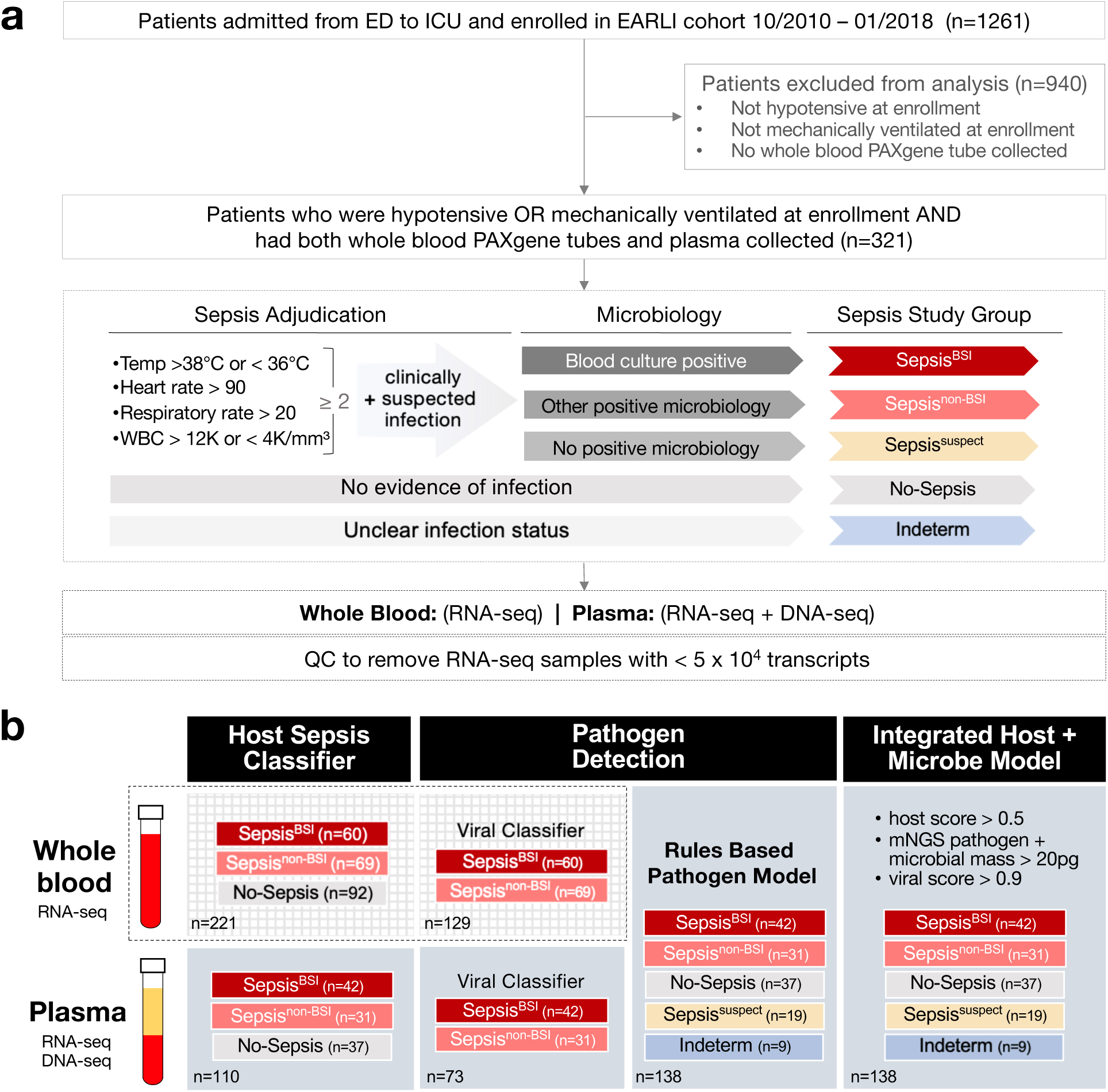
**a)** Study flow diagram. Patients studied were enrolled in the Early Assessment of Renal and Lung Injury (EARLI) cohort. Sepsis adjudication performed following hospital discharge was based on ≥ 2 or systemic inflammatory response syndrome (SIRS) criteria plus clinical suspicion of infection, and was used to delineate 5 patient subgroups. Following quality control (QC), whole blood underwent RNA-seq and plasma underwent RNA-seq and DNA-seq. **b)** Analytic approaches. Host transcriptional sepsis diagnostic classifiers were trained and tested on RNA-seq data from whole blood (n=221) and plasma (n=110), with a goal of differentiating patients with microbiologically confirmed sepsis (Sepsis^BSI^ + Sepsis^non-BSI^) from those without clinical evidence of infection (No-Sepsis). Viral infections were identified via a secondary host transcriptomic classifier. Sepsis pathogens were detected from plasma nucleic acid using metagenomic next generation sequencing (mNGS) followed by a rules-based bioinformatics model (RBM). Finally, an integrated host + microbe model for sepsis diagnosis was developed and evaluated.

### Host transcriptional signature of sepsis from whole blood

We first assessed transcriptional differences between patients with clinically and microbiologically confirmed sepsis (Sepsis^BSI^, Sepsis^non-BSI^) versus those without evidence of infection (No-Sepsis) by performing RNA sequencing (RNA-seq) on whole blood specimens (n = 221 total) to obtain a median of 5.8 x 10^7^ (95% CI 5.3-6.3 x 10^7^) reads per sample. 5,807 differentially expressed (DE) genes were identified at an adjusted P value < 0.1 (**Figure 2a, Supplementary Data 1**). Gene set enrichment analysis (GSEA), a method that identifies groups of genes within a dataset sharing common biological functions^17^, demonstrated upregulation of genes related to neutrophil degranulation and innate immune signaling in the patients with sepsis, with concomitant downregulation of pathways related to translation and rRNA processing (**Figure 2b, Supplementary Data 2**).

**Figure 2.**
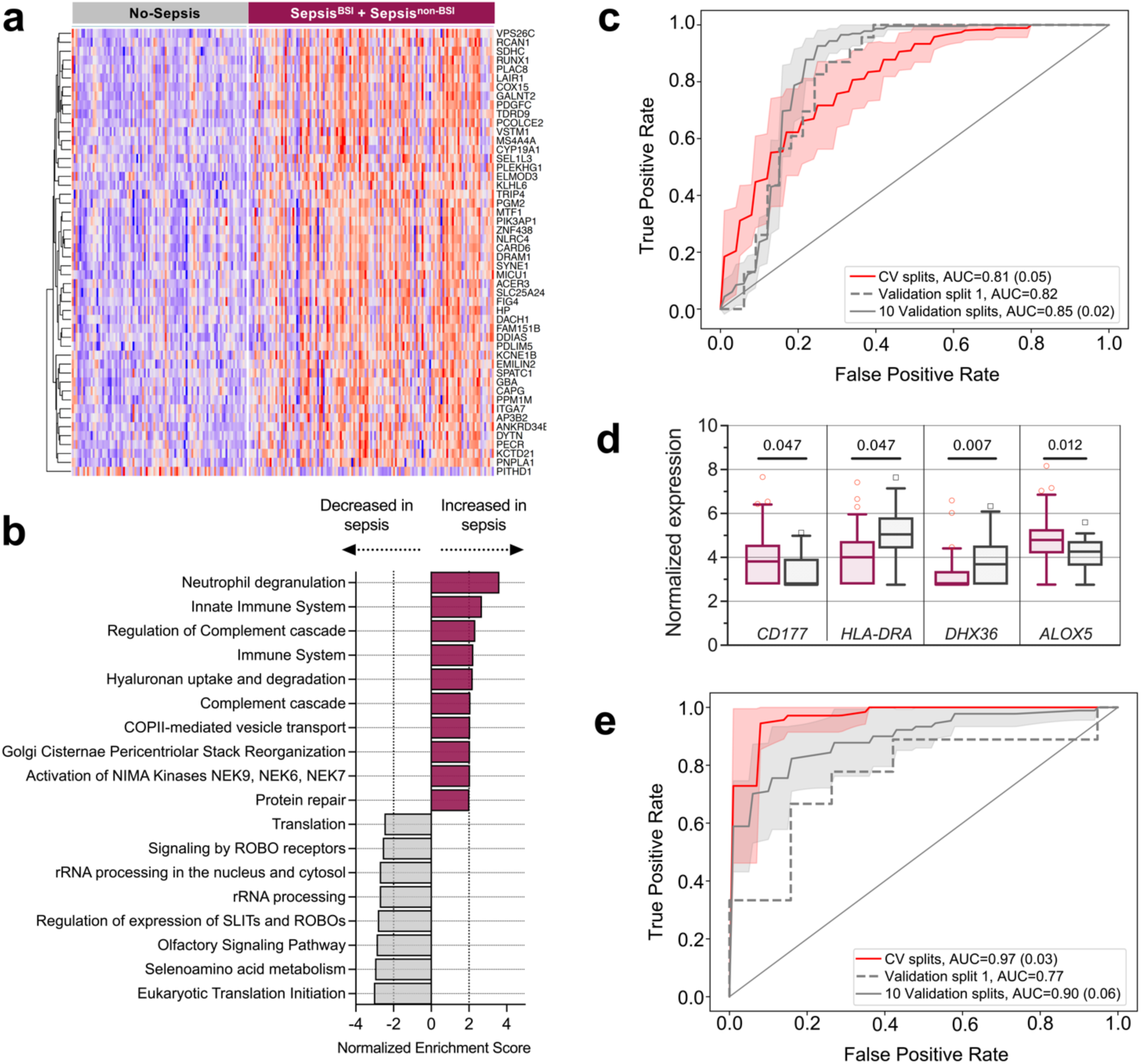
Host gene expression differentiates patients with sepsis from those with non-infectious critical illnesses. **a)** Heatmap of top 50 differentially expressed genes from whole blood transcriptomics comparing patients with microbiologically confirmed sepsis (Sepsis^BSI^ + Sepsis^non-BSI^) versus those without evidence of infection (No-Sepsis). **b)** Gene set enrichment analysis of the differentially expressed genes with the top 10 up- and down-regulated pathways (P < 0.05) highlighted. **c)** Receiver operating characteristic (ROC) curve demonstrating performance of bagged support vector machine (bSVM) classifier for sepsis diagnosis from whole blood transcriptomics (n=221). The area under the ROC curve (AUC) and standard deviation (SD, in parentheses, when applicable) are listed in the figure panel for cross validation (CV) in the training set (red line: average over 10 random splits; red shaded area: ±1SD), the held-out validation set (dashed grey line), and over 10 randomly-generated validation sets (solid grey line: average; grey shaded area: ±1SD). **d)** Plasma RNA-seq expression differences of selected differentially expressed genes previously identified as sepsis biomarkers, with Sepsis patients in maroon, and No-Sepsis patients in grey. Adjusted P value provided above boxplot. Boxes represent the 25-75 percentiles and whiskers represent the 5-95 percentiles. **e)** ROC curve demonstrating performance of bSVM classifier for sepsis diagnosis from plasma RNA (n=110). The AUC and SD are listed in the figure panel for CV in the training set (red line: average over 10 random splits; red shaded area: average ±1SD), the held-out validation set (dashed grey line), and over 10 randomly-generated validation sets (solid grey line: average; grey shaded area: average ±1SD).

To further characterize differences between sepsis patients with bloodstream versus peripheral site (e.g., respiratory, urinary tract) infections, we performed differential gene expression (DE) analysis between the Sepsis^BSI^ and Sepsis^non-BSI^ groups, which identified 5,227 genes (**Supplementary Data 3)**. GSEA demonstrated enrichment in genes related to CD28 signaling, immunoregulatory interactions between lymphoid and non-lymphoid cells, and other functions in the Sepsis^non-BSI^ patients, while the Sepsis^BSI^ group was characterized by enrichment in genes related to antimicrobial peptides, defensins, G alpha signaling and other pathways (**Supplementary Data 4**).

### Host transcriptional classifier for sepsis diagnosis from whole blood

Given the practical necessity to identify sepsis in both Sepsis^BSI^ and Sepsis^non-BSI^ patients, we constructed a ‘universal’ sepsis diagnostic classifier based on whole blood gene expression signatures. After dividing the cohort (n=221) into independent training (75% of data, n=165) and validation (25% of data, n=56) groups, we employed a bagged support vector machine learning approach (bSVM) to select genes that most effectively distinguished patients with sepsis (Sepsis^BSI^ and Sepsis^non-BSI^) from those without (No-Sepsis). We elected to use a bSVM model due to better performance compared to random forest and gradient boosted trees, which were also tested (**Supplementary Table 2**). The bSVM model achieved an average cross-validation AUC of 0.81 (standard deviation (SD) 0.05) over 10 random splits within the training dataset (75% of data, n=165). In the held-out validation set (25% of data, n=56), an AUC of 0.82 was obtained. Additionally, an AUC of 0.85 (SD 0.02) was obtained over 10 randomly-generated validation sets (**Figure 2c, Supplementary Data 5**).

### Host transcriptional classifier for sepsis diagnosis from plasma RNA

Sequencing of plasma DNA has emerged as a preferred strategy for culture-independent detection of bacterial pathogens in the bloodstream^9^. It remained unknown, however, whether plasma RNA could provide meaningful and biologically relevant gene expression data, as sepsis transcriptional profiling studies have historically relied on isolation of PBMCs or collection of whole blood.

To test this, we sequenced RNA from patients with available plasma specimens matched to the whole blood samples, and obtained a median of 2.3 x 10^7^ (95% CI 2.2-2.5 x 10^7^) reads per sample. Calculation of input RNA mass (**Methods**) demonstrated that samples with transcript counts below our QC cutoff (< 50,000) had a lower average input mass than those with sufficient counts (65.2 pg versus 85.8 pg, respectively, p <0.0001, **Supplementary Data 8**). After filtering to retain samples with ≥ 50,000 transcripts (n=138), we performed DE analysis to assess whether a biologically plausible signal could be observed between patients with sepsis (Sepsis^BSI^ and Sepsis^non-BSI^, n=73) and those without (No-Sepsis, n=37), and found 62 genes at an adjusted P value < 0.1 (**Supplementary Data 6**), 28 of which were also significant in the whole blood analysis (**Supplementary Figure 1**). Remarkably, several of the top differentially expressed genes were previously reported sepsis biomarkers (e.g., elevated *CD177,* suppressed *HLA-DRA*),^18–21^ suggesting a biologically relevant transcriptomic signature from plasma RNA (**Figure 2d, Supplementary Data 6**).

We then asked whether a host transcriptional sepsis diagnostic classifier could be constructed using plasma RNA transcriptomic data by dividing the cohort into independent training (75% of data, n=82) and validation groups (n=28), and employing the same bSVM approach to select genes that most effectively distinguished Sepsis^BSI^ and Sepsis^non-BSI^ patients from No-Sepsis patients. This approach yielded a classifier that achieved an average cross-validation AUC of 0.97 (SD 0.03) over 10 random splits within the training dataset (75% of data, n=82). In the held-out validation set (25% of data, n=28), an AUC of 0.77 was obtained. An AUC of 0.90 (SD 0.06) was obtained over 10 randomly-generated validation sets (**Figure 2e, Supplementary Data 7**).

### Detection of bacterial sepsis pathogens from plasma nucleic acid

We began microbial metagenomic analyses by assessing DNA microbial mass (**Methods**), which was significantly lower in negative control water samples, but did not differ between adjudicated sepsis groups (**Figure 3a**, **Supplementary Data 8**). We next carried out bacterial pathogen detection using the IDseq pipeline^22^ for taxonomic alignment followed by a previously developed rules-based model (RBM)^14^ that identifies established sepsis pathogens overrepresented in mNGS data compared to less abundant commensal or contaminating microbes^14^ (**Methods, Figure 3b**).

**Figure 3.**
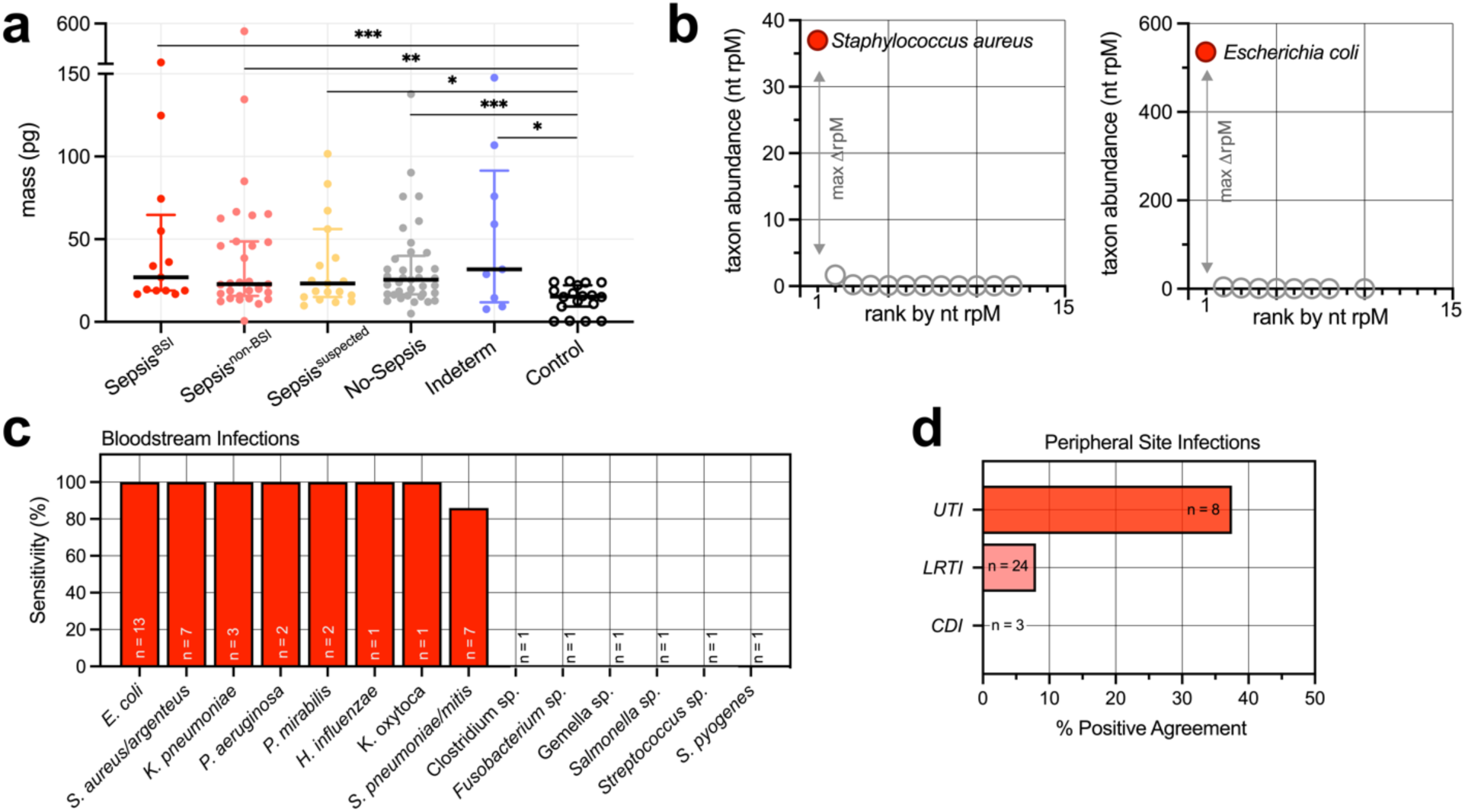
Plasma metagenomic next generation sequencing (mNGS) for detecting sepsis pathogens. **a)** Microbial plasma DNA mass differences between sepsis groups. Black bars represent median and error bars represent the interquartile range. **b)** Graphical depiction of the rules-based model (RBM) for sepsis pathogen detection from two different exemplary cases. The RBM identifies established pathogens with disproportionately high abundance compared to other commensal and environmental microbes in the sample. **c)** Concordance between plasma DNA mNGS for detecting bacterial pathogens in Sepsis^BSI^ patients with bacterial bloodstream infections compared to a gold standard of culture. **d)** Sensitivity of plasma nucleic acid mNGS for detecting pathogens in Sepsis^non-BSI^ patients with sepsis from non-bloodstream, peripheral sites of infection. Legend: LRTI = lower respiratory tract infection; UTI = urinary tract infection; CDI = *Clostridium difficile* infection. Mass data are tabulated in Supplementary Data 8. Clinical microbiology and metagenomics data are tabulated in Supplementary Data 9. *** = P ≤ 0.001, ** = P ≤ 0.01, * = P ≤ 0.05.

We then asked how well the metagenomic RBM pathogen predictions agreed with bacterial blood culture data. Polymicrobial blood cultures of ≥ 3 organisms were excluded (n=2) given their unclear clinical significance, leaving a total of 40 blood culture-positive cases available for comparison (**Supplementary Data 9**). Sensitivity versus blood culture as a reference standard was 83%, and varied by pathogen, ranging from 0% (e.g., *C. difficile*) to 100% (e.g., *E. coli, S. aureus/argenteus* **Figure 3c**). Pathogens were called by the RBM in 10/37 (27%) of patients in the No-Sepsis group, equating to a specificity of 73%.

### Detection of sepsis pathogens from peripheral sites using plasma nucleic acid

Plasma DNA mNGS identified 2/25 (8%) of culture-confirmed bacterial lower respiratory tract infection (LRTI) pathogens in the Sepsis^non-BSI^ group and 3/8 (38%) culture-confirmed bacterial urinary tract infection (UTI) pathogens (**Figure 3d, Supplementary Data 9**). mNGS did not identify *C. difficile* in any of the three patients with severe colitis from this organism. Additional putative bacterial pathogens not detected by culture were detected in 8/73 (11%) of patients with microbiologically confirmed sepsis (**Supplementary Data 9**).

### Identification of viral infections using host transcriptional profiling of RNA and whole blood

Only one of 13 (8%) respiratory viruses identified by clinical testing could be detected by plasma RNA mNGS (**Supplementary Data 9**). Recognizing that an alternative approach would be needed, we asked whether host response could instead be used to identify viral sepsis by carrying out differential gene expression analysis of patients with or without clinically confirmed viral sepsis within the Sepsis^BSI^ and Sepsis^non-BSI^ groups, using whole blood (Supplementary Data 10) or plasma (Supplementary Data 11) transcriptomic data. GSEA demonstrated that pathways related to interferon signaling and genes important for antiviral immunity were enriched in samples from patients with viral sepsis versus those with bacterial sepsis, in data derived from both whole blood (**Figure 4a, Supplementary Data 12a**) and plasma (**Figure 4b, Supplementary Data 12b**) datasets.

**Figure 4.**
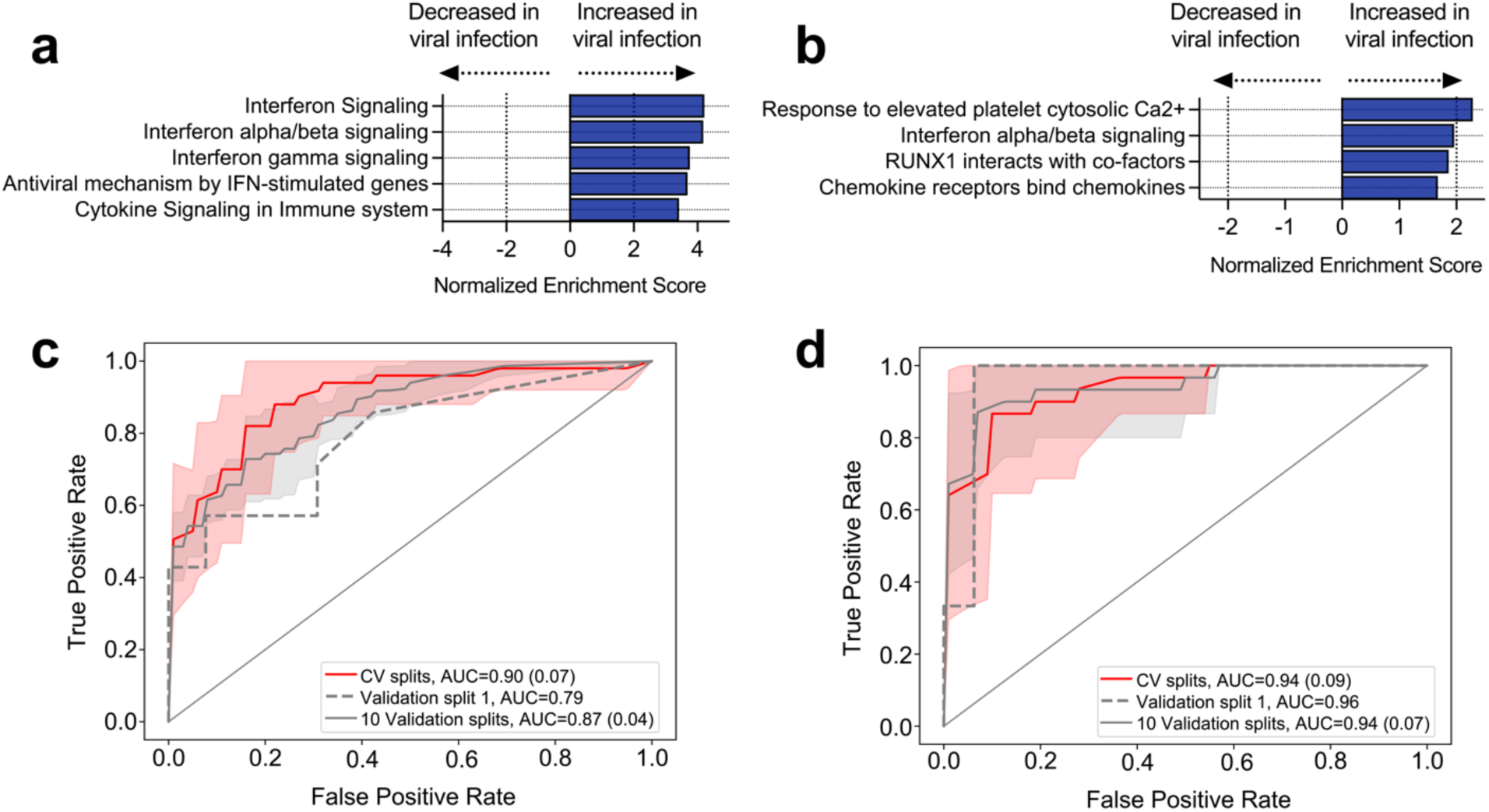
Detection of viral sepsis based on host gene expression. **a)** GSEA of differentially expressed genes from whole blood RNA-seq (n=129) demonstrating pathways enriched in patients with viral sepsis. Gene sets with P < 0.05 included. **b)** GSEA of differentially expressed genes from plasma RNA-seq (n=73) demonstrating pathways enriched in patients with viral sepsis. Gene sets with P < 0.05 included. **c)** ROC curve demonstrating performance of bagged support vector machine (bSVM) classifier for detecting viral sepsis from whole blood RNA-seq (n=129). The area under the ROC curve (AUC) and standard deviation (SD, in parentheses, when applicable) are listed in the figure panel for cross validation (CV) in the training set (red line: average over 10 random splits; red shaded area: ±1SD), the held-out validation set (dashed grey line), and over 10 randomly generated validation sets (solid grey line: average; grey shaded area: ±1SD). **d)** ROC curve demonstrating performance of bSVM classifier for detecting viral sepsis from plasma RNA-seq (n=73). The AUC and SD are listed in the figure panel for CV in the training set (red line: average over 10 random splits; red shaded area: ±1SD), the held-out validation set (dashed grey line), and over 10 randomly generated validation sets (solid grey line: average; grey shaded area: ±1SD).

We then leveraged this host signature to build a secondary bSVM diagnostic classifier for viral sepsis selecting differentially expressed genes as potential predictors, which on whole blood samples achieved an average cross-validation AUC of 0.90 (SD 0.07) over 10 random splits within the training dataset (75% of data, n=96). In the held-out validation set (25% of data, n=33), an AUC of 0.79 was obtained. An AUC of 0.87 (SD 0.04) was obtained over 10 randomly-generated validation sets (**Figure 4c, Supplementary Data 13**). Slightly better performance was obtained when building a classifier using plasma RNA-seq data, with an average cross-validation AUC of 0.94 (SD 0.09) over 10 random splits within the training dataset (75% of data, n=54). In the held-out validation set (25% of data, n=19), an AUC of 0.96 was obtained. An AUC of 0.94 (SD 0.07) was obtained over 10 randomly-generated validation sets (**Figure 4d, Supplementary Data 14**). Incorporation of the host-based viral sepsis classifier improved the sensitivity versus clinical respiratory viral PCR testing to 12/13 (92%), and predicted viral infection in one additional Sepsis^non-BSI^ patient who didn’t undergo viral PCR testing (**Supplementary Data 15**).

### Integrated host-microbe sepsis diagnostic model using plasma nucleic acid

Given the relative success of each independent host and pathogen model, we considered whether combining them could enhance diagnosis, and potentially serve as a sepsis rule-out tool. To test this possibility, we developed a proof-of-concept integrated host + microbe model based on simple rules. It returned a sepsis diagnosis based on either host criteria: `[host sepsis classifier probability > 0.5]` or microbial criteria: `[(pathogen detected by RBM) AND (microbial mass > 20 pg)] OR [host viral classifier probability > 0.9]`. Applying these rules enabled detection of 42/42 (100%) of cases in the Sepsis^BSI^ group and 30/31 (97%) of cases in the Sepsis^non-BSI^ subjects, for an overall sensitivity of 72/73 (99%) (**Figures 5a, 5b**). This proof-of-concept model yielded a specificity of 29/37 (78%) within the No-Sepsis subjects **(Figure 5c, Supplementary Data 15).**

**Figure 5.**
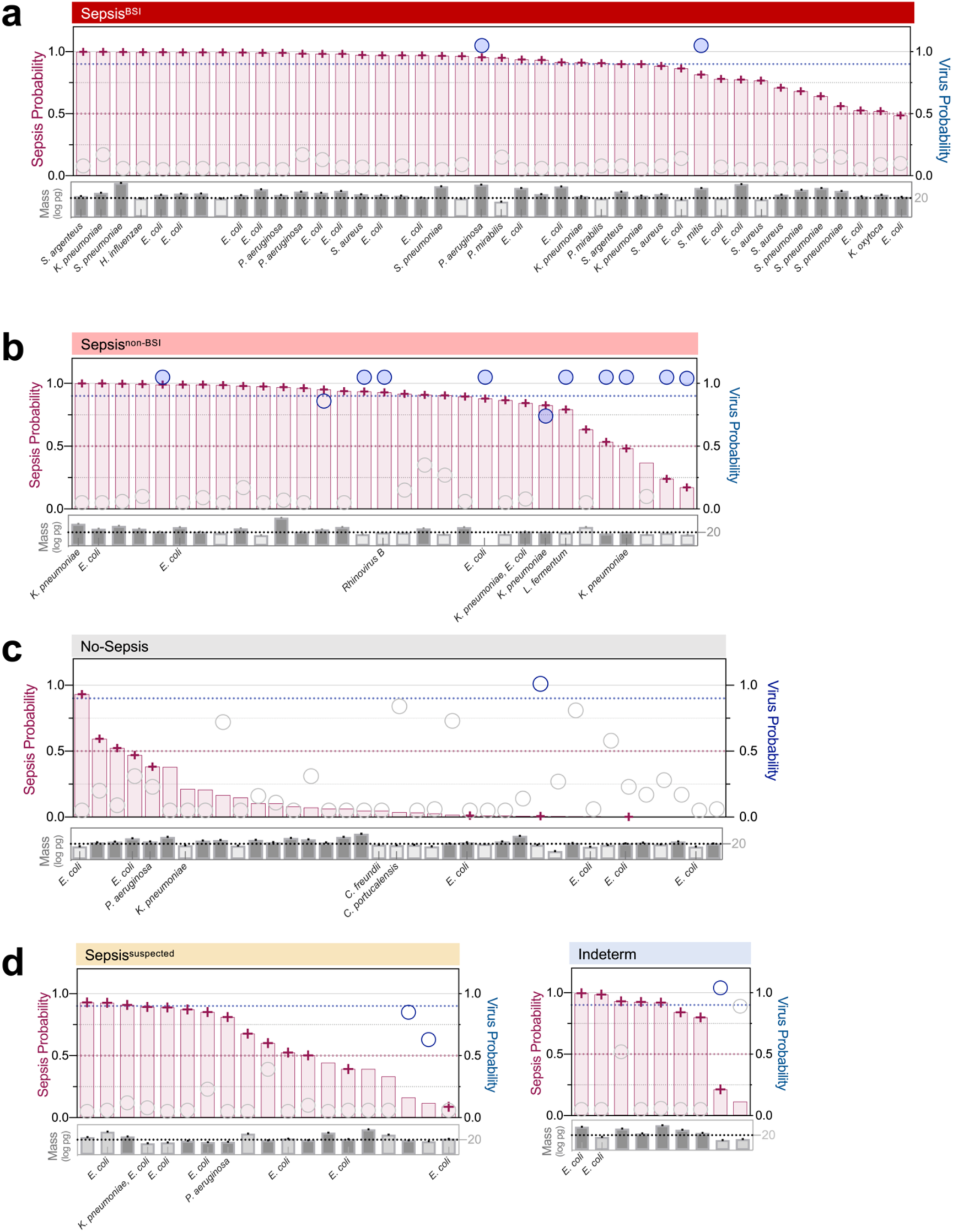
Integrated host-microbe mNGS model for sepsis diagnosis from plasma. Host criteria for positivity can be met by a sepsis transcriptomic classifier probability > 0.5 (maroon bars, dotted line). Microbial criteria can be met based on either: 1) detection of a pathogen by mNGS *and* a sample microbial mass > 20 pg (grey bars), or 2) viral transcriptomic classifier probability > 0.9 (blue circles, dotted line). Host and microbial metrics are highlighted for patients with sepsis due to **a)** bloodstream infections (Sepsis^BSI^), **b)** peripheral infection (Sepsis^non-BSI^), **c)** patients with non-infectious critical illness (No-Sepsis), and **d)** patients with suspected sepsis but negative microbiological testing (Sepsis^suspected^) and patients with indeterminant sepsis status (Indeterm). Maroon cross: sepsis positive based on model. Blue circles: virus predicted from plasma RNA secondary viral host classifier. Filled blue circles = virus also detected by clinical respiratory viral PCR. Cases with < 20pg microbial mass indicated by lighter grey shading. Samples with mNGS-detected pathogens have the microbe(s) listed below the sample microbial mass. Raw values for plots and original training/test split assignments are tabulated in Supplementary Data 16.

### Application of the integrated model to suspected and indeterminant sepsis cases

Next, we asked whether patients with clinically adjudicated sepsis, but negative in-hospital microbiologic testing (Sepsis^suspected^) would be predicted to have sepsis using the integrated host-microbe plasma mNGS model. 14/19 (74%) were classified as having sepsis, (**Figure 5d**), eight of which had a putative bacterial pathogen identified. Two additional patients had viral host classifier probabilities > 0.5, but did not meet the threshold for sepsis-positivity in the integrated model. With respect to the indeterminate group, the integrated host + microbe model classified 8/9 (89%) as sepsis-positive (**Figure 5e, Supplementary Data 15**). Of these, two had a putative bacterial pathogen identified and one had a putative viral infection identified by the viral host classifier.

### Comparison against clinical variable models for sepsis diagnosis

Lastly, we asked how host/microbe mNGS compared against sepsis diagnostic models derived exclusively from clinical metrics that would be available at the time of initial evaluation in the ED. We tested three different machine learning methods to distinguish Sepsis (Sepsis^BSI^ and Sepsis^non-BSI^) from No-Sepsis patients, using 34 clinical variables as input (**Supplementary Table 3a**). The data were split into training (75%) and validation (25%) sets, and model performance was evaluated on the latter. The greatest average AUC achieved was 0.62 (SD 0.04) using a random forest model (**Supplementary Table 3b**). We then computed the AUC using the qSOFA score, a widely used clinical score for sepsis diagnosis^15^. The qSOFA achieved an average AUC of 0.48 (SD 0.02).

## Discussion

Sepsis is defined as a dysregulated host response to infection^15^, yet existing diagnostics have focused exclusively on either detecting pathogens or assessing features of the infected host. Here, we combined host transcriptional profiling with broad-range pathogen detection to accurately diagnose sepsis in critically ill patients upon hospital admission. Further, we demonstrate that an integrated host-microbe metagenomics approach can be performed on circulating RNA and DNA from plasma, a widely available clinical specimen type with previously unrecognized utility for host-based infectious disease diagnosis.

Identifying an etiologic pathogen is critical for optimal treatment of sepsis. We found that concordance between pathogen detection by plasma mNGS and traditional bacterial blood culture varied by organism. For instance, mNGS sensitivity for detecting *S. aureus* and *E. coli*, two of the most globally important sepsis pathogens^5^, was 100%. In contrast, mNGS missed several important but less common sepsis pathogens, such as *S. pyogenes*. We noted that in all false-negative cases, the patients had received antibiotics prior to mNGS sample collection, which may have reduced the abundance of circulating bacterial DNA available for detection. Furthermore, mNGS and blood cultures were performed on different samples, with research specimens collected up to 24 hours after blood cultures, which may have resulted in lower concordance than if samples had been collected contemporaneously.

Several of the microbes missed by mNGS were organisms that in many contexts exist as commensals (e.g., Fusobacterium, Gemella and Streptococcus species). It is unclear whether these organisms were truly etiologic sepsis pathogens or commensals translocated to the blood in the setting of critical illness, and incidentally identified in culture. Clostridium species are frequently found as contaminants in blood cultures^23^, which is perhaps why mNGS also did not detect an unspeciated Clostridium isolated in blood culture from one patient.

With respect to non-BSI sepsis, our findings suggest that plasma mNGS may be most useful for identifying UTI-associated pathogens, although we also observed some utility for respiratory pathogen detection, in line with a prior report^24^. mNGS failed to detect *C. difficile* in any patients with colitis from this pathogen, although this is not surprising given that the organism is rarely associated with bacteremia^25^.

Within the No-Sepsis group, 10/37 (27%) of patients had a pathogen detected by mNGS, each of which was validated to be present at statistically significantly higher levels than in background controls. Notably, 9/10 (90%) of the pathogens were gram negative enteric organisms, which may reflect gastrointestinal translocation of microbes, a well described phenomenon during critical illness^26^. In addition, all 10 of these patients had received antibiotics in the first day of study enrollment, so it is possible that sequences derived from non-viable organisms unable to grow in culture.

Plasma RNA sequencing alone performed poorly for detecting sepsis-associated respiratory viruses. Incorporation of a host-based viral classifier, however, markedly improved detection of clinically confirmed viral LRTI. The viral classifier predicted previously unrecognized viral infections in three patients with sepsis who did not undergo viral PCR testing during their hospitalizations. Prior work has demonstrated that different viral species elicit distinct host transcriptional signatures in the peripheral blood^27^, suggesting that future studies could extend the RNA host viral classifier to identify specific viral pathogens, such as influenza or SARS-CoV-2, for which therapeutics exist. Future studies could additionally explore the use of targeted enrichment methods^28, 29^ to enhance detection of viral sepsis pathogen nucleic acid.

In line with prior reports^13^, we found that viral sepsis has a unique host transcriptional signature characterized by expression of interferon and other signaling pathways. We also observed transcriptional differences based on whether sepsis was due to a bloodstream versus peripheral site infection, which was less expected. The host response in Sepsis^BSI^ patients was characterized by lower expression of genes related to CD28 signaling and T cell activation, and greater expression of genes related to antimicrobial peptides, defensins and G alpha signaling, compared to Sepsis^non-BSI^ patients.

We found that detection of a pathogen alone was in many cases insufficient for sepsis diagnosis, but when combined with a host transcriptional profile, had promising diagnostic utility and potential as a tool for infection rule-out. In addition to defining host signatures of sepsis from whole blood, we also found biologically relevant host transcripts in plasma. This may have direct clinical applications given that plasma mNGS is increasingly being used in hospitals for pathogen detection in patients with sepsis and other infectious diseases, with turnaround times of ≤48 hours.

Inappropriate antimicrobial use is a major challenge in the management of critical illness, and is often driven by the inability to rule-out infection in patients with systemic inflammatory diseases. Indeed, we found that clinical variables alone, including the qSOFA score, were unable to accurately distinguish patients with sepsis from those with non-infectious critical illnesses at the time of initial evaluation in the ED. In contrast, our proof-of-concept assessment of the integrated host + microbe mNGS model demonstrated 99% sensitivity across patients with microbiologically confirmed sepsis, and 78% specificity within the No-Sepsis group, which was comprised almost entirely of patients meeting the clinical definition of systemic inflammatory response syndrome^16^.

Host/microbe mNGS may facilitate precision antimicrobial stewardship by discriminating sepsis from diverse types of non-infectious febrile inflammatory syndromes, ranging from autoimmune diseases to macrophage activation syndrome. We envision this assay being used at the time of ED presentation for all suspected sepsis patients, as an adjunct to blood cultures and other traditional microbiological testing.

Distinguishing true sepsis pathogens from environmental contaminants or human commensals (e.g., Gemella), is a challenge for both mNGS and traditional culture-based microbiologic methods. Concomitant assessment of a host-based metric offers an opportunity to determine whether the detected pathogen exists in the context of an immunological state consistent with infection. Considering this, host/microbe mNGS diagnostic classification could theoretically be more difficult in immunocompromised patients. Arguing against this, however, is prior work demonstrating accurate performance of a host/microbe mNGS pneumonia diagnostic in an ICU cohort with a 40% prevalence of immunocompromised individuals^14^.

Our study has several strengths, including the novel use of plasma RNA transcriptomics for sepsis diagnosis, development of the first sepsis diagnostic combining host and microbial mNGS data, detailed clinical phenotyping, and a large prospective cohort of critically ill adults with systemic illnesses. It also has some limitations. First, as noted above, mNGS and blood cultures were performed on different samples collected at different times, so the observed concordance with clinical microbiological testing may be an underestimate. Second, a significant fraction of plasma samples had insufficient host transcripts to permit gene expression analyses, leading to a smaller sample size for the plasma versus the whole blood cohorts. This limitation may be addressable in future studies by increasing the input plasma volume and thus RNA mass.

The host immune response during sepsis is dynamic, and thus the stage of infection at which gene expression is measured may influence accuracy of the classifier. While our study was cross-sectional in design, we attempted to control for this by sampling at a consistently early stage of critical illness, within the first 24 hours of ICU admission. Lastly, because we did not have access to any other sepsis studies with either plasma gene expression data or paired host and microbial mNGS data from blood, additional studies in an independent cohort will be needed to validate these findings.

In conclusion, we report that combining host gene expression profiling and metagenomic pathogen detection from plasma nucleic acid enables accurate diagnosis of sepsis. Future studies are needed to validate and test the clinical impact of this culture-independent diagnostic approach.

## Methods

### Study design, clinical cohort, and ethics statement

We conducted a prospective observational study of patients with acute critical illnesses admitted from the ED to the ICU. We studied patients who were enrolled in the Early Assessment of Renal and Lung Injury (EARLI) cohort at the University of California, San Francisco (UCSF) or Zuckerberg San Francisco General Hospital between 10/2010 and 01/2018 (**Supplementary Table 1**). The study was approved by the UCSF Institutional Review Board (IRB) under protocol 10-02852, which granted a waiver of initial consent for blood sampling. Informed consent was subsequently obtained from patients or their surrogates for continued study participation, as previously described^30, 31^.

For the parent EARLI cohort, the inclusion criteria are: 1) age ≥ 18, 2) admission to the ICU from the ED, and 3) enrollment in the ED or within the first 24 hours of ICU admission. For this study, we selected patients for whom PAXgene whole blood tubes and matched plasma samples from the time of enrollment were available. PAXgene tubes were collected on patients enrolled in EARLI during the time period listed above who were hypotensive and/or mechanically ventilated at the time of enrollment. The main exclusion criteria for the EARLI study are: 1) exclusively neurological, neurosurgical, or trauma surgery admission, 2) goals of care decision for exclusively comfort measures, 3) known pregnancy, 4) legal status of prisoner, and 5) anticipated ICU length of stay < 24 hours. Enrollment in EARLI began in 10/2008 and continues.

### Sepsis adjudication

Clinical adjudication of sepsis groups was carried out by study team physicians (MA, CL, AL, KL, PS, CH, AG, CC, KK, MM) using the sepsis-2 definition^32^ (≥ 2 SIRS criteria + suspected infection) and incorporating all available clinical and microbiologic data from the entire ICU admission, with blinding to mNGS results. Each patient was reviewed by at least four physicians. Disagreements were handled by discussion with the most senior physicians (CC, MM) in the phenotyping panel. Patients were categorized into five subgroups based on sepsis status (**Figure 1a, Supplementary Figure 1**). Patients with clinically adjudicated sepsis and a bacterial culture-confirmed bloodstream infection (Sepsis^BSI^), sepsis due to a microbiologically confirmed primary infection at a peripheral site other than the bloodstream (Sepsis^non-BSI^), suspected sepsis with negative clinical microbiologic testing (Sepsis^suspected^), patients with no evidence of sepsis and a clear alternative explanation for their critical illness (No-Sepsis), or patients of indeterminant status (Indeterm). Clinical and demographic features of patients are summarized in (**Supplementary Table 1**) and tabulated in (**Supplementary Data 16 and 17**).

### Metagenomic sequencing

Following enrollment, whole blood and plasma were collected in PAXgene and EDTA tubes, respectively. Whole blood PAXgene tubes were processed and stored at −80C according to manufacturer’s instructions, and plasma was frozen at −80C within two hours. To evaluate host gene expression and detect microbes, RNA-seq was performed on the whole blood and plasma specimens, and DNA-seq was performed on plasma specimens. RNA was extracted from whole blood using the Qiagen RNeasy kit and normalized to 10ng total input per sample. Total plasma nucleic acid was extracted by first clarifying 300uL of plasma via maximum-speed centrifugation for five minutes at 21,300 x g, and then employing the Zymo Pathogen Magbead Kit on the supernatant following manufacturer’s instructions. 10ng of total nucleic acid underwent DNA-seq using the NEBNext Ultra II DNA Kit. Samples with at least 10ng of remaining total nucleic acid were treated with DNAse (Qiagen) to recover RNA, and then underwent RNA-seq library preparation using the NEBNext Ultra II RNA-seq Kit as described below.

For RNA-seq library preparation, human cytosolic and mitochondrial ribosomal RNA and globin RNA was first depleted using FastSelect (Qiagen). For the purposes of background contamination correction (see below) and to enable estimation of input microbial mass, we included negative water controls as well as positive controls (spike-in RNA standards from the External RNA Controls Consortium (ERCC))^33^. RNA was then fragmented and underwent library preparation using the NEBNext Ultra II RNA-seq Kit (New England Biolabs) according to described methods . Finished libraries underwent 146 nucleotide paired-end Illumina sequencing on an Illumina Novaseq 6000 instrument.

Index swapping can lead to read misassignment with Illumina sequencing. Dual indexing, that is adding barcode index sequences on both ends of the molecule, reduces the rate at which this misassignment occurs by requiring concordance between the two barcode sequences. The frequency of index-swapped reads has been estimated to be more than 35X lower when using dual vs single indexing^35^. Because we used dual indexing and because the RBM for pathogen detection operates by only identifying pathogen sequences disproportionately abundant in a sample versus the other sequences, our methods would not be expected to be negatively influenced by index swapping, which would only be anticipated to misassign low abundance reads irrelevant to the RBM.

### Host differential expression and pathway analysis

Following demultiplexing, sequencing reads were aligned with STAR^36^ to an index consisting of all transcripts associated with human protein coding genes (ENSEMBL v. 99), cytosolic and mitochondrial ribosomal RNA sequences, and the sequences of ERCC RNA standards. Samples retained in the dataset had a total of at least 50,000 counts associated with transcripts of protein coding genes.

Differential expression analysis was performed using DESeq2 and including covariates for age and gender. Significant genes were identified using an independent-hypothesis-weighted, Benjamini-Hochberg false discovery rate (FDR) < 0.1^38, 39^. We generated heatmaps of the top 50 differentially expressed genes by absolute log_2_-fold change. To evaluate signaling pathways from gene expression data, we employed gene set enrichment analysis using WebGestalt^40^ on all ranked differentially expressed genes with a P value < 0.1. Significant pathways and upstream regulators were defined as those with a gene set P value < 0.05.

### Pathogen detection

Detection of microbes leveraged the open-source IDseq pipeline^22^ which incorporates subtractive alignment of the human genome (NCBI GRC h38) using STAR^36^, quality and complexity filtering, and subsequent removal of cloning vectors and phiX phage using Bowtie2^22^. The identities of the remaining microbial reads are determined by querying the NCBI nucleotide (NT) database using GSNAP-L^22, 41^. After background correction (see below), retained non-viral taxonomic alignments in each sample were aggregated at the genus level, and sorted in descending order by abundance measured in reads per million (rpM), independently for each sample. A previously validated rules based model (RBM)^14^ was then utilized to identify disproportionately abundant bacteria and fungi in each sample, and flag them as pathogens. The RBM, originally developed to identify pathogens from respiratory mNGS data, detects outlier organisms within a sample by identifying the greatest gap in abundance between the top 15 sequentially ranked microbes in each sample. All microbes present in a reference index of established pathogens above this gap are then called by the RBM.

We adapted the original RBM specifically for sepsis pathogen detection, in which outlier organisms are sometimes present in low abundance, by incorporating a sepsis (as opposed to respiratory) pathogen reference index (**Supplementary Data 18**) and requiring that the species called by the RBM both be present in the reference index and detected at an abundance of > 1 rpM. Given the potential for respiratory viruses to cause sepsis, the RBM also identified human pathogenic respiratory viruses derived from a reference list of LRTI pathogens^14^, present in the plasma RNA-seq data at an abundance of > 1 rpM. Sensitivity and specificity were calculated based on detection of reference index sepsis pathogens in each of the sepsis adjudication groups.

The reference index (**Supplementary Data 18**) was established *a. priori* and no data from the enrolled patients were used to inform the distinction between pathogens and commensals. The index consisted of the most prevalent bloodstream infection pathogens reported by both the National Healthcare Safety Network (NHSN)^42^ and a recent multicenter surveillance study of healthcare-associated infections^43^. These studies reported multiple species of Bacteriodes, Candida, Citrobacter, Enterobacter, Enterococcus, Klebsiella, Lactobacillus, Morganella, Prevotella, Proteus, Serratia, Stenotrophomonas and Streptococcus as common sepsis pathogens, and thus the reference index contains all species within these genera, yielding > 1000 total species detectable by the model based on current NCBI taxonomy.

### Identification and mitigation of environmental contaminants

Negative control samples consisting of only double-distilled water (n=24) were processed alongside plasma DNA samples, which were sequenced in a single batch. Negative control samples enabled estimation of the number of background reads expected for each taxon^44^. A previously developed negative binomial model^44^ was employed to identify taxa with NT sequencing alignments present at an abundance significantly greater compared to negative water controls. This was done by modeling the number of background reads as a negative binomial distribution, with mean and dispersion fitted on the negative controls. For each taxon, we estimated the mean parameter of the negative binomial by averaging the read counts across all negative controls. We estimated a single dispersion parameter across all taxa, using the functions glm.nb() and theta.md() from the R package MASS^45^. Taxa that achieved an adjusted P value <0.01 (Benjamini & Hochberg multiple test correction) were carried forward to the above-described RBM for pathogen detection.

### Microbial mass calculations

Microbial mass was calculated based on the ratio of microbial reads in each sample to total reads aligning to the External RNA Controls Consortium (ERCC) RNA standards spiked into each sample^46^. The following equation was utilized for this calculation: [ERCC input mass]/[microbial input mass] = [ERCC reads]/[microbial reads], where the ERCC input mass was 25pg.

### Host transcriptional classifiers for sepsis and viral infection diagnosis

To build classifiers that differentiated patients with sepsis (Sepsis^BSI^, Sepsis^non-BSI^) from those with non-infectious critical illness (No-Sepsis), and distinguished viral from non-viral sepsis, we built a Support Vector Machine (SVM)-based classifier^47^ with the scikit-learn^48^ (v0.23.2) library in Python (v3.8.3). We tested several machine learning approaches (bagged SVM, random forest and gradient boosted trees) and selected a bSVM classifier with a linear kernel based on best performance (**Supplementary Table 2**). Each classifier used a bootstrapped set of samples and a random subset of features.

We evaluated samples with ≥ 50,000 plasma gene counts and genes with more than 20% non-zero counts in that sample subset. Only differentially expressed genes, identified using DESeq2 (v1.28.1) in the training set, were considered as potential predictors and included in machine learning models, with FDR thresholds of 0.1 (whole blood), 0.2 (plasma, viral) and 0.3 (plasma, sepsis) chosen based on cross-validation. Age and sex were included as covariates in the models. We used Z-score-scaled transformed (variance stabilizing transformation) gene counts. 75% of the data was selected to train the model, and the rest was used as a held-out set to test the final model. The training set was subsequently randomly split ten times for cross-validation, using 75% of each as intermediate training sets, and the remaining 25% as their associated testing sets.

On each one of those intermediate training sets, we carried out feature selection and parameters optimization using nested 5-fold cross-validations. We optimized three parameters: the regularization parameter, the maximum number of features considered for each classifier, and the total number of classifiers to use for bagging. For each parameters optimization fold, a recursive feature elimination (RFE) strategy was adopted, dropping 10% of the remaining least important features at each iteration. A bSVM classifier with default parameters was built at each iteration. We defined feature importance as the average squared weight across all estimators. To maximize interpretability, we restricted the maximum number of predictors to 100 genes.

We estimated model performances using the Area Under the Receiver Operating Characteristic Curve (AUC) values. To obtain a single set of features, we fitted a model, using the aforementioned strategy, to the initial training set. This model was then tested on the held-out set to obtain a final performance value and a single set of predictors.

### Comparison of plasma nucleic acid mNGS against clinician-ordered diagnostic testing

Clinical microbiological testing was carried out based on decisions from the primary medical team during the patient’s hospital admission at the UCSF and ZSFG clinical microbiology laboratories Tests utilized included bacterial culture from blood, lower respiratory tract and urine which were carried out according to previously described protocols^14^. Clinical testing for viral respiratory pathogens was performed from nasopharyngeal swabs and/or bronchioalveolar lavage using the Luminex XTag multiplex viral PCR assay. Polymicrobial blood cultures with ≥ 3 bacteria (n=2) were excluded from pathogen concordance given their unclear clinical significance and potential that some organisms reflected contamination.

### Integrated host + microbe sepsis diagnosis and rule-out model

We developed a simple integrated host + microbe model that returned a sepsis diagnosis based on either host criteria `[host sepsis classifier probability > 0.5]` or microbial criteria: `[(pathogen detected by RBM) AND (microbial mass > 20 pg)] OR [host viral classifier probability > 0.9]`. Combined metrics (**Supplementary Data 16**) including sepsis assignment based on this model are depicted in Figure 5. Sensitivity was calculated in the (Sepsis^BSI^) and (Sepsis^non-BSI^) groups, and specificity in the (No-Sepsis) group.

### Clinical variable models for sepsis diagnosis

We tested the ability of clinical variables (**Supplementary Table 3a**) available at the time of initial patient assessment to predict sepsis using three machine learning methods. These included SVM using the e1071 package^49^, random forest using the randomForest package^50^ and regularized logistic regression using the glmnet^51^ package in R version 4.2.0^52^. Specifically, we built models to classify Sepsis (Sepsis^BSI^ and Sepsis^non-BSI^) versus No-Sepsis using 34 clinical variables that would be available at the time of ED evaluation. The data were split into training (75%) and test (25%) sets and model performance (AUC) was evaluated on the test set. This was repeated for a total of 10 randomized splits with the AUC computed at each iteration. AUC was also computed for the qSOFA score (systolic blood pressure < 100 mmHg, respiratory rate > 22 breaths/minute, Glasgow Coma Scale < 13). Results are tabulated in (**Supplementary Table 3b**).

### Statistics and reproducibility

Statistical tests utilized for each analysis are described in the figure legends and in further detail in each respective methods section. The number of patient samples analyzed for each comparison are indicated in the figure legends. Data were generated from single sequencing runs without technical replicates.

### Data availability

Source data are provided with this paper. The processed genecount data are available from the National Center for Biotechnology Information Gene Expression Omnibus database under accession code GSE189403. The raw sequencing data are protected due to data privacy restrictions from the IRB protocol governing patient enrollment, which protects the release of raw genetic sequencing data from those patients enrolled under a waiver of consent. To honor this, researchers who wish to obtain raw fastq files for the purposes of independently generating genecounts can contact the corresponding author and request to be added to the IRB protocol. The raw fastq files with microbial sequencing reads are available from the Sequence Read Archive under BioProject ID: PRJNA783060.

### Code availability

Code for the differential expression, classifier development and RBM can be found at: (https://github.com/lucile-n/plasma_classifiers).

## Data Availability

The processed genecount data are available from the National Center for Biotechnology Information Gene Expression Omnibus database under accession code GSE189403. The raw fastq files with microbial sequencing reads are available from the Sequence Read Archive under BioProject ID: PRJNA783060.

## Supplementary Materials

**Supplementary Table 1a.**
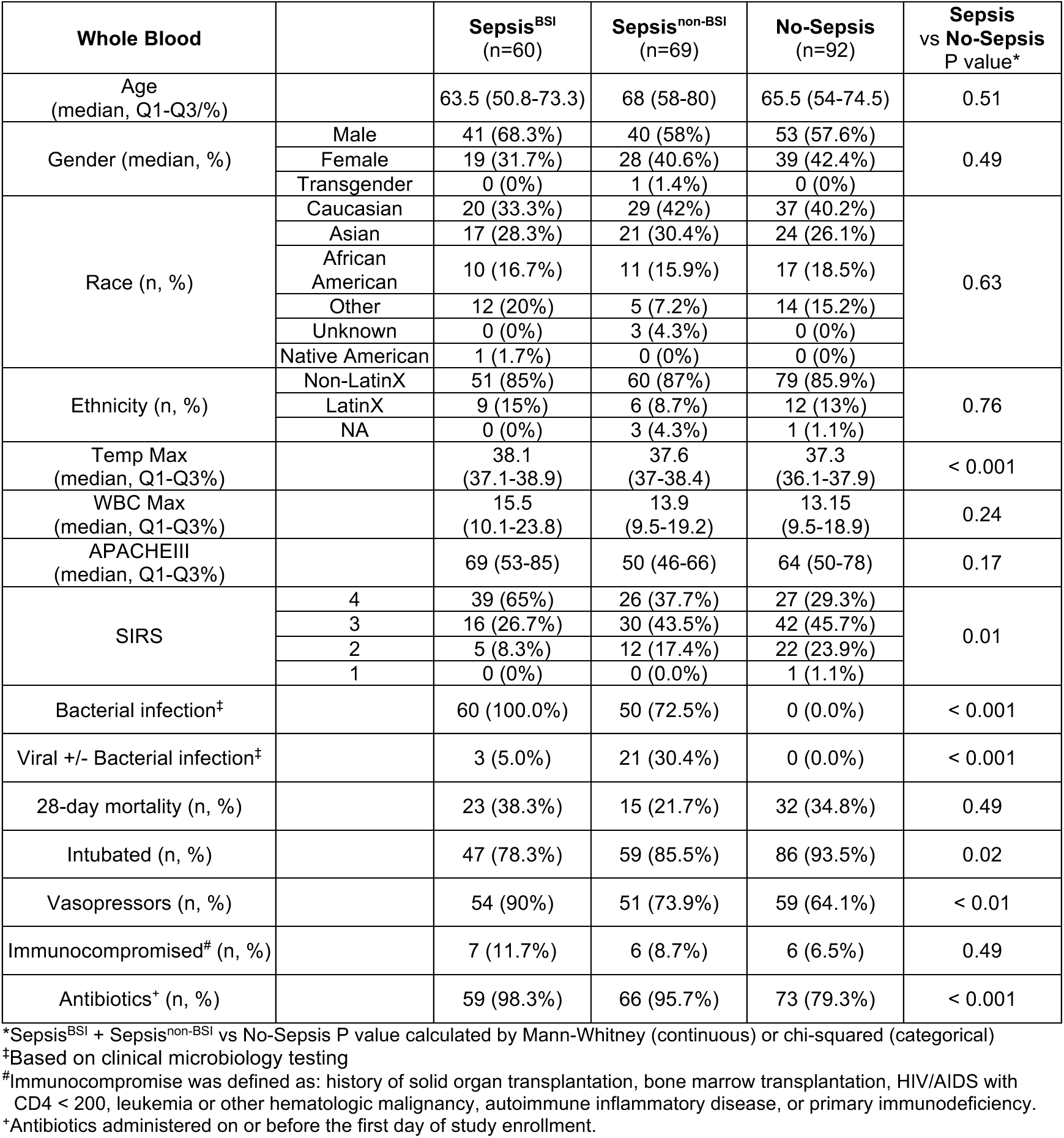
Summary of clinical and demographic features of patients evaluated in whole blood gene expression analyses (n=221). These include patients with microbiologically confirmed sepsis (Sepsis^BSI^ and Sepsis^non-BSI^) and those with non-infectious critical illnesses (No-Sepsis). Source data are tabulated in (Supplementary Data 16).

**Supplementary Table 1b.**
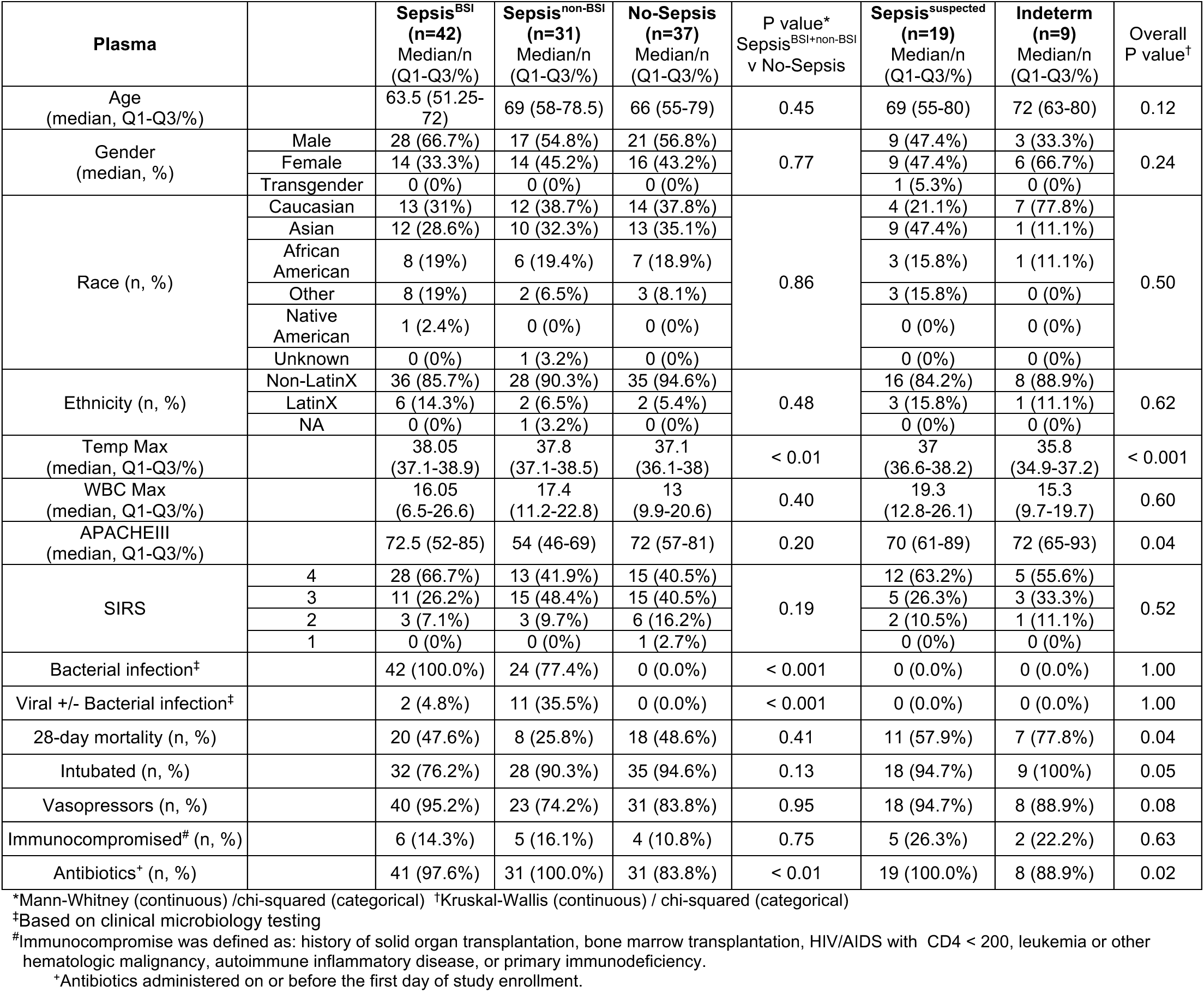
Summary of clinical and demographic features of patients with plasma RNA-seq data, evaluated in all analyses (n=138). All sepsis adjudication groups represented. Source data are tabulated in (Supplementary Data 17).

**Supplementary Table 2.** **Comparison of machine learning models for host-based sepsis classification.** Area under the receiver operator characteristic curve (AUC) for three different machine learning models assessed for classifier construction. The AUC for cross-validation in the training set (standard deviation in parentheses) is listed first, and the AUC for the first validation split is listed second, after the vertical bar. The bagged support vector machine (bSVM) model performed best overall.

| Classifier | Bagged Support Vector Machine | Random Forest | Gradient Boosted Tree |
| --- | --- | --- | --- |
| Whole blood - sepsis | 0.81 (0.05) \| 0.82 | 0.80 (0.06) \| 0.86 | 0.79 (0.05) \| 0.84 |
| Plasma - sepsis | 0.97 (0.03) \| 0.77 | 0.76 (0.05) \| 0.79 | 0.70 (0.09) \| 0.82 |
| Whole blood - viral | 0.90 (0.07) \| 0.79 | 0.83 (0.08) \| 0.75 | 0.78 (0.07) \| 0.69 |
| Plasma - viral | 0.94 (0.09) \| 0.96 | 0.77 (0.09) \| 0.81 | 0.72 (0.19) \| 0.60 |

**Supplementary Table 3a.**
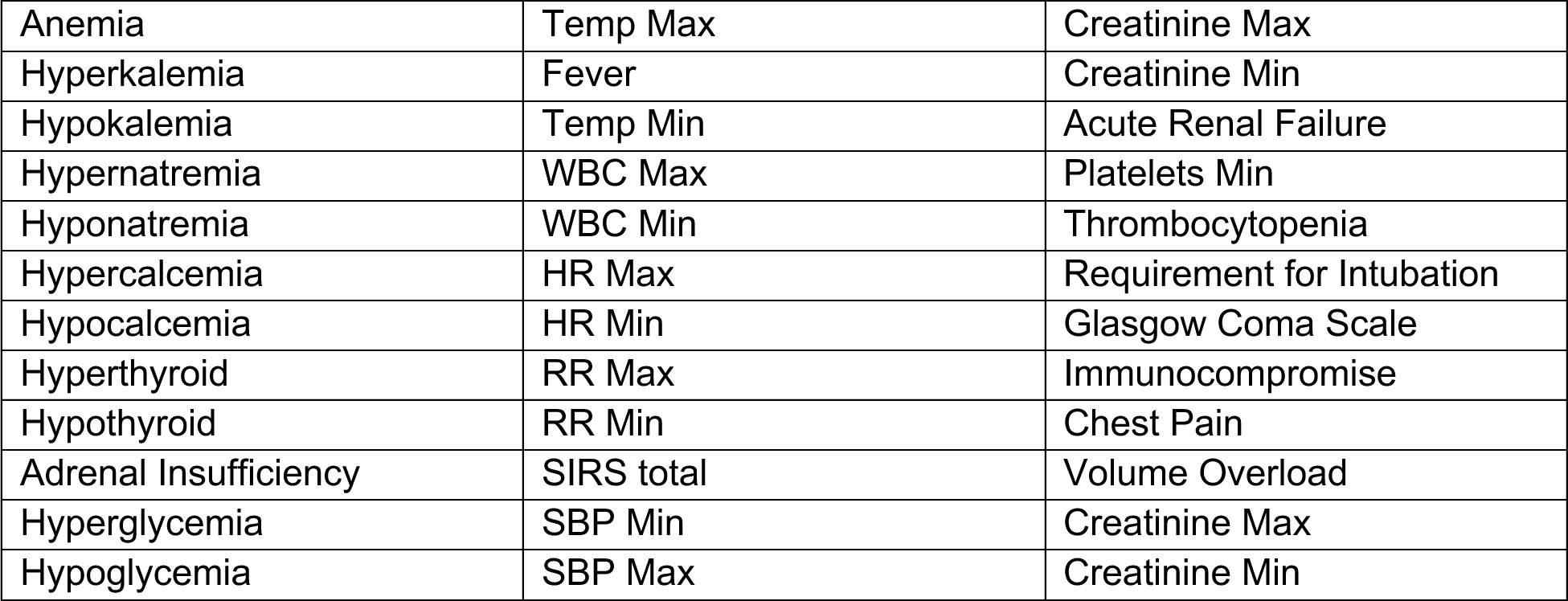
Clinical variables used for classifier construction. Fever: > 38C, anemia: hemoglobin < 7, thrombocytopenia: platelets < 50, acute renal failure: creatinine > 2.0.

**Supplementary Table 3b.**
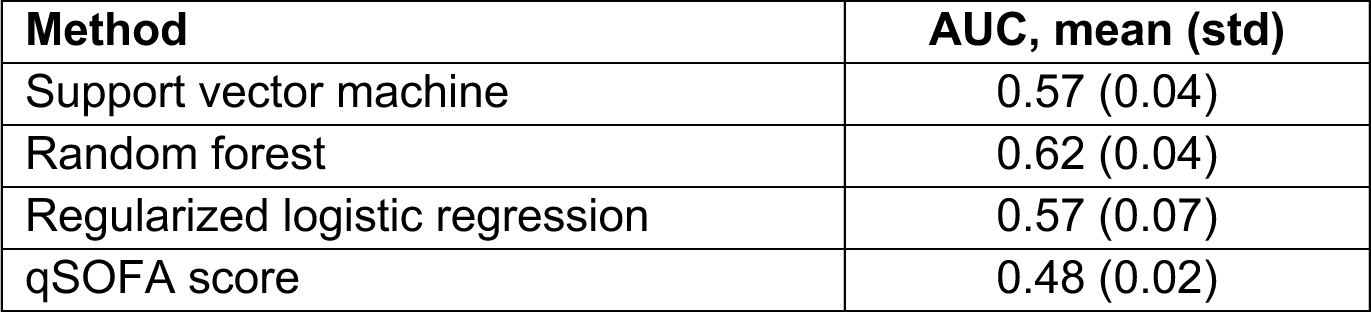
Average classifier AUC over 10 iterations of training and test from different machine learning classifiers to distinguish Sepsis from No-Sepsis patients using clinical features alone. qSOFA score positive for sepsis if systolic blood pressure < 100 mmHg, respiratory rate > 22 breaths/minute and Glasgow Coma Scale < 13.

## Supplementary Figures

**Supplementary Figure 1.**
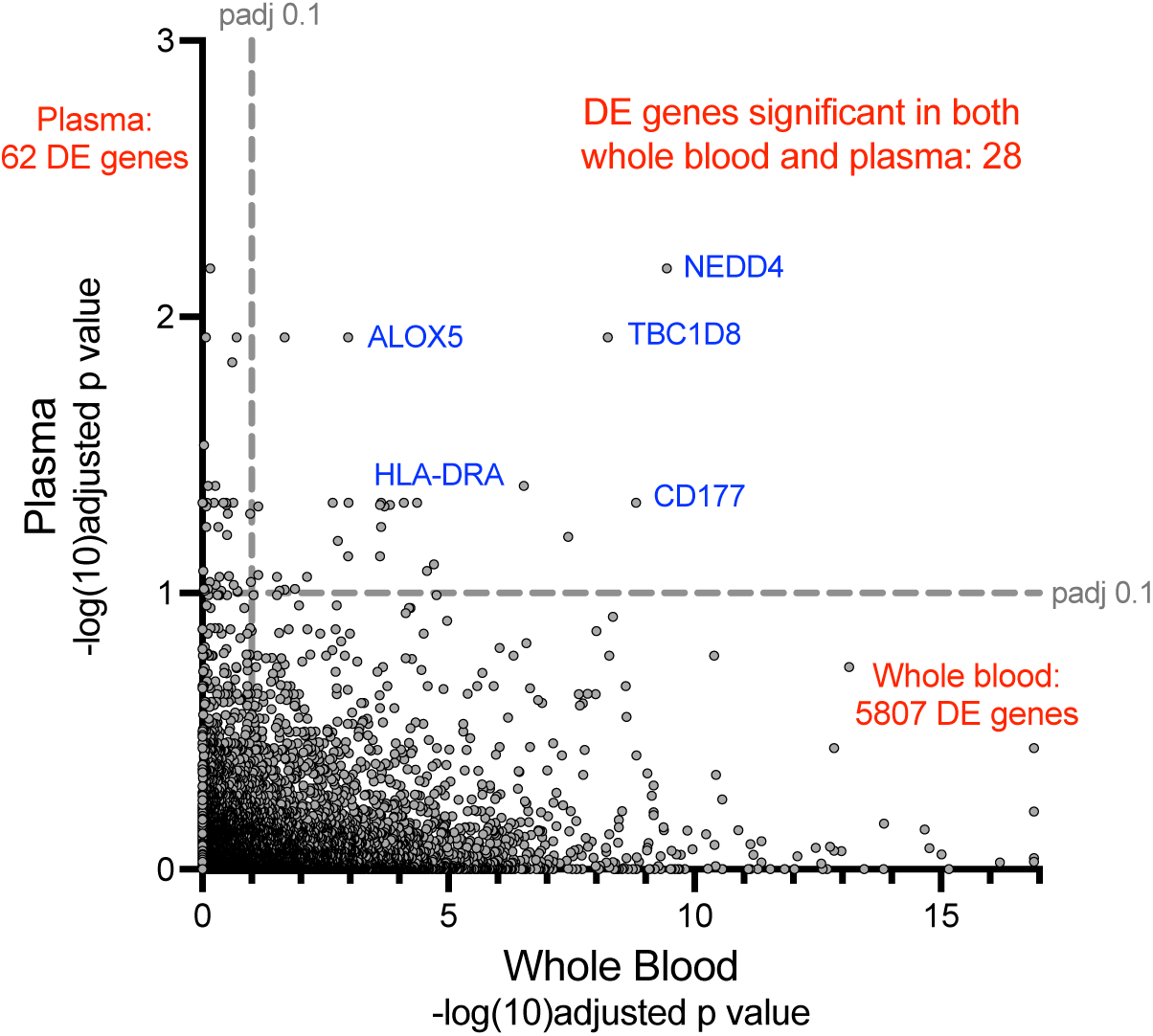
Overlap of significant genes in the differential expression analyses between the Sepsis and No-Sepsis groups for whole blood and plasma samples. Scatter plot of −log10(adjusted p-value) for individual genes from the differential expression analyses comparing patients with microbiologically confirmed sepsis (Sepsis^BSI^ + Sepsis^non-BSI^) versus those without evidence of infection (No-sepsis), from whole blood (x-axis) and plasma (y-axis). P-values derive from Benjamini-Hochberg adjustment. Dashed gray lines indicate the threshold of adjusted p-value < 0.1. Selected, significant, differentially expressed genes highlighted in blue.

**Supplementary Figure 2.**
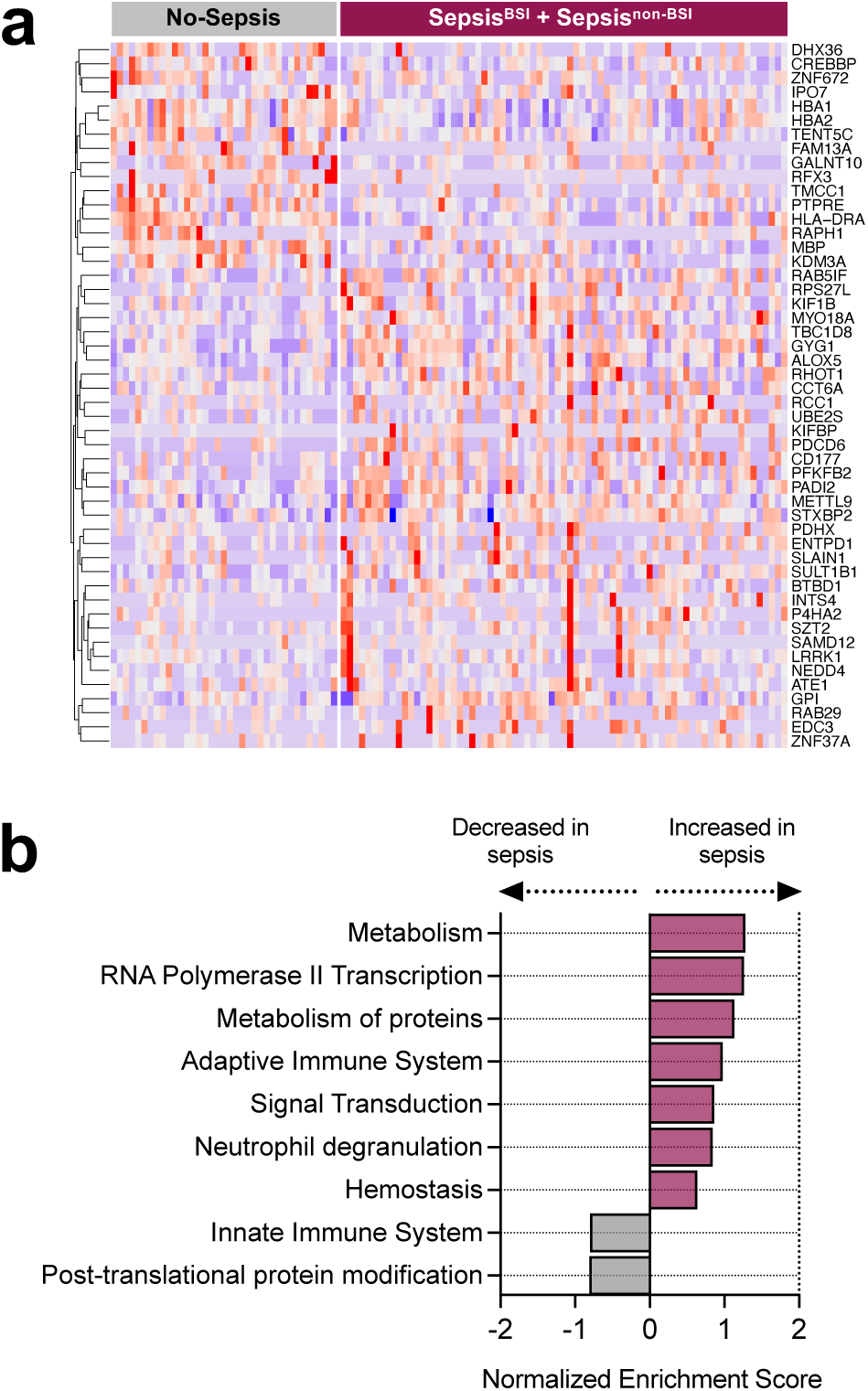
Plasma host gene expression differentiates patients with sepsis from those with non-infectious critical illnesses. **a)** Heatmap of top 50 differentially expressed genes from whole blood transcriptomics comparing patients with microbiologically confirmed sepsis (Sepsis^BSI^ + Sepsis^non-BSI^) versus those without evidence of infection (No-sepsis). **b)** Gene set enrichment analysis of the differentially expressed genes. All gene sets included.

## Supplementary Data Files

Supplementary Data 1. Differentially expressed genes (adjusted P value < 0.1) between patients with microbiologically confirmed (Sepsis^BSI^ and Sepsis^non-BSI^) and those with non-infectious critical illnesses (No-Sepsis), from whole blood RNA-seq.

Supplementary Data 2. Gene set enrichment analysis of differentially expressed genes between patients with microbiologically confirmed sepsis (Sepsis^BSI^ and Sepsis^non-BSI^) and those with non-infectious critical illnesses (No-Sepsis). Data from whole blood RNA-seq. The top 10 positively and negatively enriched pathways by P value are included in table.

Supplementary Data 3. Differentially expressed genes (adjusted P value < 0.1) between patients with sepsis due to bloodstream infections (Sepsis^BSI^) versus peripheral infections (Sepsis^non-BSI^). Data from whole blood RNA-seq.

Supplementary Data 4. Gene set enrichment analysis of differentially expressed genes between patients with sepsis due to bloodstream infections (Sepsis^BSI^) versus peripheral infections (Sepsis^non-BSI^). Data from a) whole blood RNA-seq and b) plasma RNA-seq. The top 10 positively and negatively enriched pathways by P value are included in table.

Supplementary Data 5. a) Area under the receiver operating characteristic curve (AUC) values for 10 independent training set models for a whole blood gene expression support vector machine classifier to distinguish patients with microbiologically confirmed (Sepsis^BSI^ and Sepsis^non-BSI^) from those with non-infectious critical illnesses (No-Sepsis). b) Composite list of all genes selected by each classifier model.

Supplementary Data 6. Differentially expressed genes (adjusted P value < 0.1) between patients with microbiologically confirmed (Sepsis^BSI^ and Sepsis^non-BSI^) and those with non-infectious critical illnesses (No-Sepsis), from plasma RNA-seq.

Supplementary Data 7. a) AUC values for 10 independent training set models for a plasma gene expression support vector machine classifier to distinguish patients with microbiologically confirmed (Sepsis^BSI^ and Sepsis^non-BSI^) from those with non-infectious critical illnesses (No-Sepsis). b) Composite list of all genes selected by each classifier model.

Supplementary Data 8. Mass (pg) of microbial DNA in each sample, calculated based on spiked-in 25 pg ERCC positive controls.

Supplementary Data 9. Sepsis pathogens detected by standard of care clinical microbiology versus plasma mNGS, using the rules-based model.

Supplementary Data 10. Differentially expressed genes (adjusted P value < 0.1) between patients with microbiologically confirmed viral sepsis and those with non-viral sepsis (Sepsis^BSI^ and Sepsis^non-BSI^ groups), from whole blood RNA-seq.

Supplementary Data 11. Differentially expressed genes (adjusted P value < 0.1) between patients with microbiologically confirmed viral sepsis and those with non-viral sepsis (Sepsis^BSI^ and Sepsis^non-BSI^ groups), from plasma RNA-seq.

Supplementary Data 12. Gene set enrichment analysis of differentially expressed genes between patients with viral versus non-viral causes of sepsis amongst the Sepsis^BSI^ and Sepsis^non-BSI^ patients. a) Data from whole blood RNA-seq. b) Data from plasma RNA-seq.

Supplementary Data 13. a) AUC values for 10 independent training set models for a whole blood gene expression support vector machine classifier to distinguish patients with microbiologically confirmed viral versus non-viral sepsis (Sepsis^BSI^ and Sepsis^non-BSI^), from whole blood RNA-seq. b) Composite list of all genes selected by each classifier model.

Supplementary Data 14. a) AUC values for 10 independent training set models for a plasma gene expression support vector machine classifier to distinguish patients with microbiologically confirmed viral versus non-viral sepsis (Sepsis^BSI^ and Sepsis^non-BSI^), from plasma RNA-seq. b) Composite list of all genes selected by each classifier model.

Supplementary Data 15. Complete integrated host-microbe mNGS dataset. This includes: per-sample classifier predictions for all patients with plasma sequencing data (n=138), including the sepsis diagnostic classifier and the viral sepsis classifier; pathogens detected by clinical diagnostics and by mNGS; and microbial mass per sample.

Supplementary Data 16. Clinical and demographic features of patients evaluated in whole blood gene expression analyses only (n=221). These include patients with microbiologically confirmed sepsis (Sepsis^BSI^ and Sepsis^non-BSI^) and those with (No-Sepsis).

Supplementary Data 17. Clinical and demographic features of patients with plasma RNA-seq data, evaluated in all analyses (n=138). All sepsis adjudication groups represented.

Supplementary Data 18. Reference index of established sepsis pathogens derived from the top 20 most prevalent sepsis pathogens reported by both the US CDC/ National Healthcare Safety Network^1^ and a point prevalence survey of healthcare-associated infections^2^.

